# Plasma proteomics and coronary artery calcium score: synergistic, concordant and contrasting predictions of cardiovascular outcomes in The Multi-Ethnic Study of Atherosclerosis

**DOI:** 10.64898/2026.05.12.26353070

**Authors:** Jessica Chadwick, Meredith Carpenter, Matthew Budoff, Rajat Deo, Ruth F. Dubin, Philip Greenland, Michael A. Hinterberg, Rajeev Malhotra, Clint Miller, Jerome I. Rotter, Kent D. Taylor, Emma V. Troth, Peter Ganz

## Abstract

**Background:** Coronary artery calcium (CAC) scores inform subclinical atherosclerotic cardiovascular disease (ASCVD) burden, helping guide preventative treatments. However, prediction of cardiovascular (CV) events by CAC is largely limited to ASCVD outcomes. This study investigated whether a previously validated proteomic test for predicting a broad composite of four-year CV events could enhance the prognostic utility of CAC.

**Methods:** We used a 27-protein CV risk score (Prot-CVR), derived from ∼5,000 SomaScan™ Assay plasma protein measurements, to predict four-year risk of a composite CV and mortality outcome (myocardial infarction, stroke/TIA, heart failure hospitalization, death) in 2,122 participants with ≥1 CV risk factors from the Multi-Ethnic Study of Atherosclerosis (MESA) observational cohort at exam 5 and compared predictions to CAC Agatston scores. Discriminatory performance was assessed using C-Index and 4-year area under the curve (AUC). Cox Proportional Hazard (CoxPH) ratios were calculated for the composite outcome, ASCVD outcome (myocardial infarction, resuscitated cardiac arrest, stroke, coronary heart disease death), and individual events. Changes in Prot-CVR and CAC scores from baseline to MESA exam 5 (+10-years) in CV event versus event-free participants were assessed using 2-tailed paired t-tests. CoxPH regression models of CV event status distributed by Prot-CVR, CAC, and relevant co-variates were evaluated for performance relative to individual models.

**Results:** Individual Prot-CVR and CAC models predicting the composite outcome had comparable 4-year AUCs, but Prot-CVR had a higher C-index (0.68 (0.65-0.70) versus 0.63 (0.60-0.65), p=0.001) and greater hazard ratios for the composite outcome (p<0.001), death (p<0.001), and heart failure (p=0.015). A combined CoxPH model of Prot-CVR + CAC + Age had a higher 4-year AUC (0.72, p<0.05) and C-Index (0.71, p<0.05) than Prot-CVR or CAC alone. Both Prot-CVR and CAC scores detected an increase in risk prior to an approaching CV event in ∼10-year sensitivity-to-change analysis. For 49.6% of MESA population with CAC=0 at baseline, Prot-CVR was greater in composite event versus event free participants at 4 years (0.23 versus 0.15, p=0.006) and full follow-up (0.18 versus 0.13, p<0.001).

**Conclusion:** Protein testing complements CAC for CV risk assessment although the improvement is modest. Prot-CVR may resolve which patients with CAC=0 are at heightened CV risk.

## Introduction

Coronary artery calcium (CAC) scores are a powerful measure of subclinical coronary atherosclerosis burden that can guide the need for lipid-modifying therapy and other atherosclerotic cardiovascular disease (ASCVD) preventive measures in asymptomatic individuals. CAC scores have utility primarily as strong predictors of coronary event risk, including myocardial infarction (MI)^1, 2^ although they have also been shown to be associated with cerebrovascular events^3, 4^, incident heart failure^5, 6^, and all-cause mortality^7^. While CAC scores are used as a risk assessment tool in the general population, they have also been applied for risk assessment in specific high-risk individuals including those with type 2 diabetes^8^, chronic kidney disease^9^, suspected coronary artery disease^10^ and the elderly^11^.

However, limitations of CAC scores include inability to rule out clinically significant noncalcified atherosclerotic plaques, particularly in patients with a CAC score = 0, where it is estimated 4-7% have non-calcified plaques. CAC Agatston scores also have substantial interscan variability notably at the low end of the scores, ranging from 15% to 22% due to noise, motion, type of scanner, reconstruction window and partial volume averaging, all of which can lead to overestimation of the calcium score^12^. Furthermore, statins can paradoxically increase CAC scores as they enhance calcification of existing plaques and increase plaque density as part of the stabilization process^13^.

Non-imaging-based risk assessment tools, such as blood-based proteomics, could complement and augment the prognostic utility of CAC and enhance clinical risk stratification. We previously developed and validated a 27-protein model (Prot-CVR) in a total of 22,849 participants in nine clinical studies that predicts four-year risk for a CV event – defined as the first occurrence of MI, stroke/transient ischemic attack (TIA), heart failure (HF) hospitalization, or all-cause death – among individuals with heightened CV risk^14^. This 27-protein risk score predicts CV risk with greater discrimination, broader dynamic range, and better risk reclassification than the ACC/AHA Pooled Cohort ASCVD risk score^14^.

The goal of the present analyses was to evaluate how Prot-CVR compares to CAC score in a heightened risk and asymptomatic population (participants with ≥ 1 known causes of increased CV risk, including individuals with stable cardiovascular disease (CVD), type 2 diabetes, chronic kidney disease, a history of cancer, symptoms consistent with chronic coronary syndromes, or age > 65 years) using the Multiethnic Study of Atherosclerosis (MESA) baseline and exam 5 (+10-years) cohort data, and determine whether Prot-CVR scoring can serve as a complementary tool to CAC scores.

## Methods

### Study population

MESA is a study of the characteristics of subclinical cardiovascular disease and the risk factors that predict progression to clinically overt cardiovascular disease or progression of the subclinical disease^15^. MESA consists of a diverse, community-based sample of an initial 6,814 men and women aged 45-84 years without known cardiovascular disease at baseline. Thirty-eight percent of the recruited participants were White, 28% African American, 22% Hispanic, and 12% of Chinese descent. Participants were recruited from six field centers across the United States: Baltimore City and Baltimore County, Maryland; Chicago, Illinois; Forsyth County, North Carolina; Los Angeles County, California; New York, New York; and St. Paul, Minnesota. Participants were followed for identification and characterization of cardiovascular disease events, including acute myocardial infarction and other forms of coronary heart disease (CHD), stroke, and heart failure; for cardiovascular disease interventions; and for mortality. The first examination took place over two years, from July 2000 to July 2002, and has been followed by additional examinations. Participants were contacted every 9 to 12 months throughout the study to assess and adjudicate clinical morbidity and mortality. The study was approved by the institutional review boards at all participating institutions, and all participants gave written informed consent. In addition, informed consent was obtained for extensive data sharing (dbGaP) and genetic/omic studies, including candidate genes (NHLBI CARe), genome-wide scans (NHLBI SHARe), exome sequencing (NHLBI ESP) and, most recently, the NHLBI TOPMed program. A detailed description of the study design and methods has been published previously^15^.

Participants were excluded from the proteomics analysis if incident CV event follow-up information or date of censoring was missing, CAC Agatston data were not available, participants were not part of the proteomic CV risk test intended use population (defined as individuals ≥40 years with one or more known causes of increased CV risk, including individuals with: stable CVD, type 2 diabetes, chronic kidney disease, a history of cancer, or age > 65 years) or their plasma samples did not meet standard QC and pre-analytic requirements for the SomaScan Assay (n=876). Fully informed written consent for participation in the study and consent to the use of data for research were obtained from all evaluable participants, and the study was reviewed and approved by the local ethics committees at participating MESA study sites. At that time of the consent procedure, 11% of otherwise eligible MESA participants opted not to have their blood samples analyzed by a commercial company (in this case SomaLogic) and were excluded from these analyses.

### Proteomic profiling

The modified aptamer binding reagents^16^, SomaScan v4.1 Assay^17, 18^, and its performance characteristics^19^ have been previously described. The assay uses DNA-based slow off-rate modified aptamers (SOMAmer™ Reagents) to quantify the relative abundance of approximately 7,000 proteins with high specificity and limits of detection largely comparable to antibody-based assays. Briefly, the SomaScan Assay begins as a mixture of ∼7,000 SOMAmer Reagents labelled with a 5’ fluorophore, photocleavable linker, and biotin, which are immobilized on streptavidin (SA)-coated beads and incubated with 55 µL of EDTA plasma samples. A polyanionic competitor is added to prevent non-specific interactions between SOMAmer Reagents and non-cognate protein targets. SOMAmer Reagents are hybridized to complementary sequences on a DNA microarray chip and quantified by fluorescence, with signal intensity proportional to the number of bound SOMAmer Reagents and, by extension, the relative abundance of the target protein. This enables quantitative comparison of differential protein expression across samples and time points. Standard normalization procedures were conducted^20^.

### Proteomic cardiovascular risk test

The development and validation of 27-protein CV risk test (Prot-CVR, formally known as The Residual Cardiovascular Risk – 4 years SomaSignal™ Test) utilized in these analyses has been described previously.^14^ The protein variables and their weighting coefficients are shown in Supplementary Table S1. The test predicts the likelihood of a CV event (defined as a MI, stroke/TIA, HF hospitalization or all-cause death) within 4 years in individuals ≥40 years of age with one or more known causes of elevated CV risk, including individuals with stable CVD (history of MI or stroke (>6 months prior), HF, peripheral artery disease, revascularization, abnormal stress test or imaging suggesting coronary heart disease), type 2 diabetes, chronic kidney disease, a history of cancer, symptoms consistent with chronic coronary syndromes, or age over 65 years old. The test was developed and validated on the SomaScan v4.0 (∼5,000 protein) Assay using 32,130 plasma samples from 22,849 participants across multiple studies including: The Trøndelag Health Study, The Atherosclerosis Risk in Communities (ARIC) Study, The BASEL VIII Study, The Exenatide Study of Cardiovascular Event Lowering Trial, and The Chronic Renal Insufficiency Cohort. Prot-CVR was subsequently validated on the (∼7,000 protein) SomaScan v4.1 Assay, the version of the platform used to assay the MESA samples. The test reports results as an absolute risk score and relative risk category.

Predictive performance comparisons and combined model analyses utilized stored plasma samples and clinical data from MESA exam 5 (∼10 years after the baseline exam) since it had the largest number of participants with Agatston CAC scores that were part of the Prot-CVR intended use population. Longitudinal analyses [i.e., risk score trajectory demonstrating sensitivity to detect a change in predicted risk in participants having (or not) a subsequent outcome event] utilized data from participants with paired exam 1 (baseline) and exam 5 samples.

### Performance of the Prot-CVR score compared to CAC (Agatston score) in predicting composite cardiovascular outcomes and individual event types

The prognostic performance of Prot-CVR and CAC Agatston scores were assessed as continuous measures at predicting two separate outcomes: (1) a composite CV outcome defined as a MI, stroke/TIA, HF hospitalization or all-cause death, and (2) an ASCVD outcome defined as a MI, resuscitated cardiac arrest, stroke, coronary heart disease (CHD) death, or stroke death. Time-to-event and time-to-censoring data were used in conjunction with predicted Prot-CVR and CAC scores, and event status (event or event-free during specified follow-up period) to generate Receiver Operating Characteristic (ROC) curves, Area Under the Curve (AUC) performance metrics for 4-year censored data, and Concordance-Index (C-Index) for uncensored time-to-event data over the full follow-up (∼10 years). For the 4-year censoring, participants were classified as “event” if they had a qualifying event within the 4-year period, “no event” if they did not have a qualifying event within the 4-year period and were coded as “NA” and censored from the 4-year analysis if they did not have an event and their follow-up was less than 4 years. Delong’s test was used to compare the AUC values for ROC curves generated for Prot-CVR and CAC scores for each group. A Z-score test was constructed by the compareC (version 1.3.2)^1^ package to compare the sets of C-Indices and determine significant differences in metric performance with right-censored survival outcomes^21^.

Kaplan-Meier (KM) survival probabilities were used to evaluate the Prot-CVR and CAC scores as categorical measures at predicting the Prot-CVR validated outcome. KM survival plots were generated to visualize the time-to-event data, allowing for the comparison of survival distributions between different risk groups. Prot-CVR relative risk bins used the following categorical cutoffs that we defined as low risk (≤ 7.5%), medium-low risk (>7.5% and ≤ 25%), medium-high risk (>25% and ≤ 50%), and high risk (>50%). CAC scores were partitioned according to established guidelines^22^ as no detectable calcium (CAC = 0), mild calcification (CAC = 1-100), moderate calcification (CAC = 100-300), and extensive calcification (CAC = 300+). Additionally, the relative risk of the composite CV outcome for Prot-CVR and CAC were compared. Relative risk was calculated separately for each risk bin as 4-year event rate / background event rate, where the 4-year event rate is defined as number of CV events within 4-years / total number of individuals in the risk category and the background event rate is defined as number of people with CV events in 4-year censored data / total number of people in the 4-year censored data.

Cox PH Ratios (HR) were calculated for two outcomes (CV and mortality composite outcome and ASCVD outcome), as well as the individual event types included in one or both combined composite outcomes. CoxPH models were generated for each evaluated outcome using follow-up days to event or censoring and event status, distributed by center-scaled metric (Prot-CVR or CAC) as:

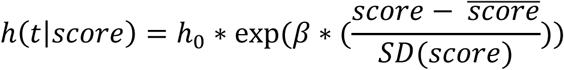

where *h(t|score)* is the hazard at time *t* for a given value of the score (Prot-CVR or CAC), *h_0_(t)* is the baseline hazard, and β is multiplied by the center scaled score. Thus, the coefficient β represents the log hazard ratio for a 1 standard deviation (SD) increase in the score; to interpret this coefficient on the hazard scale it is exponentiated as Hazard ratio = exp(β)^23^. A z-statistic calculated from the difference in log hazard ratio divided by the standard error of the difference was used to evaluate the significance of the difference between Prot-CVR and CAC Hazard Ratios for each event type. A false discovery rate (FDR) adjustment was applied to p-values to correct for multiple testing.

### Performance of a combined Prot-CVR + CAC model and adjustment for traditional risk factors

A CoxPH survival model was generated for the composite CV outcome distributed by center scaled Prot-CVR and adjusted in an add-one manner for clinical covariates known to impact cardiovascular risk [Diabetes status, sex, high-density lipoprotein cholesterol (mg/dL), systolic blood pressure (mmHg), smoking status, total cholesterol (mg/dL)], ethnicity, antihypertensive medication use, and body mass index (BMI), and for center-scaled CAC. Separately, two CoxPH combined models (Prot-CVR+CAC only and Prot-CVR+CAC+AGE) were generated to compare combined model performance to individual metric performance calculated previously. A paired Delong’s test was used to compare 4-year AUC of the models, and the compareC Z-score was used for C-Index comparisons.

### Sensitivity to detect change in risk as measured by Prot-CVR score compared to CAC in advance of an upcoming CV event

Assessment of the change in Prot-CVR predictions and CAC scores over time was evaluated on paired longitudinal samples from MESA exams 1 and 5 (∼10 years apart) to predict outcome events after exam 5. The analysis was stratified by event status: individuals who had an event (using the CV and mortality composite outcome) after exam 5 versus participants who did not have an event during the full ∼10-year follow-up period. Differences in Prot-CVR and CAC scores were calculated and evaluated for significance using 2-tailed t-tests with alpha=0.05: exam 1 (baseline) mean scores of participants who had an event were compared to those who did not have an event during the full follow-up period; mean within subject (paired samples) change in scores from exam 1 to exam 5 were compared in participants who had an event versus those who did not have an event during the full follow-up period. A sub-cohort analysis was also conducted on participants with non-zero CAC scores (CAC > 0), using paired samples from participants with non-zero CAC at both exam 1 and exam 5.

### Cardiovascular risk stratification using Prot-CVR in participants with CAC = 0

To evaluate the predictive performance of Prot-CVR in individuals with no detectable calcification (CAC = 0), the Prot-CVR scores in the sub cohort of individuals with CAC = 0 at Exam 1 were compared between the event (using CV and mortality composite outcome with 4-year censoring and over the full follow-up period) and “no event” groups with a 1-tailed t-test with the alternative hypothesis that the mean difference (event mean Prot-CVR – no event mean Prot-CVR) is greater than 0 (alpha = 0.05).

## RESULTS

### Prognostic performance of individual risk scores

Prognostic performances of Prot-CVR and CAC were assessed in 2,122 individuals from MESA exam 5. For the composite CV and mortality outcome, the overall event rates were 9.73% and 25.9% over 4 years and the full 10-year follow-up period, respectively. Characteristics of the MESA participants used for these analyses are described in Table 1 and Supplementary Table S2. Prot-CVR and CAC had a C-Index of 0.68 and 0.63 (p = 0.001) over the full time-to-event data, and a 4-year AUC of 0.67 and 0.66 (p = 0.649) in predicting the CV and mortality composite outcome, respectively (Table 2). The median time to event was 2.73 years. Both Prot-CVR and CAC performed comparably at predicting 4-year events, with comparable sensitivity and specificity demonstrated in the ROC curves (Supplemental Figure S1A). However, Prot-CVR predictive performance, sensitivity, and specificity improved over the full 10-year follow-up period while CAC predictive performance diminished, leading to a significant difference in C-indices (p = 0.001) and greater separation in ROC curves for the two scores (Supplemental Figure S1B).

**Table 1:**
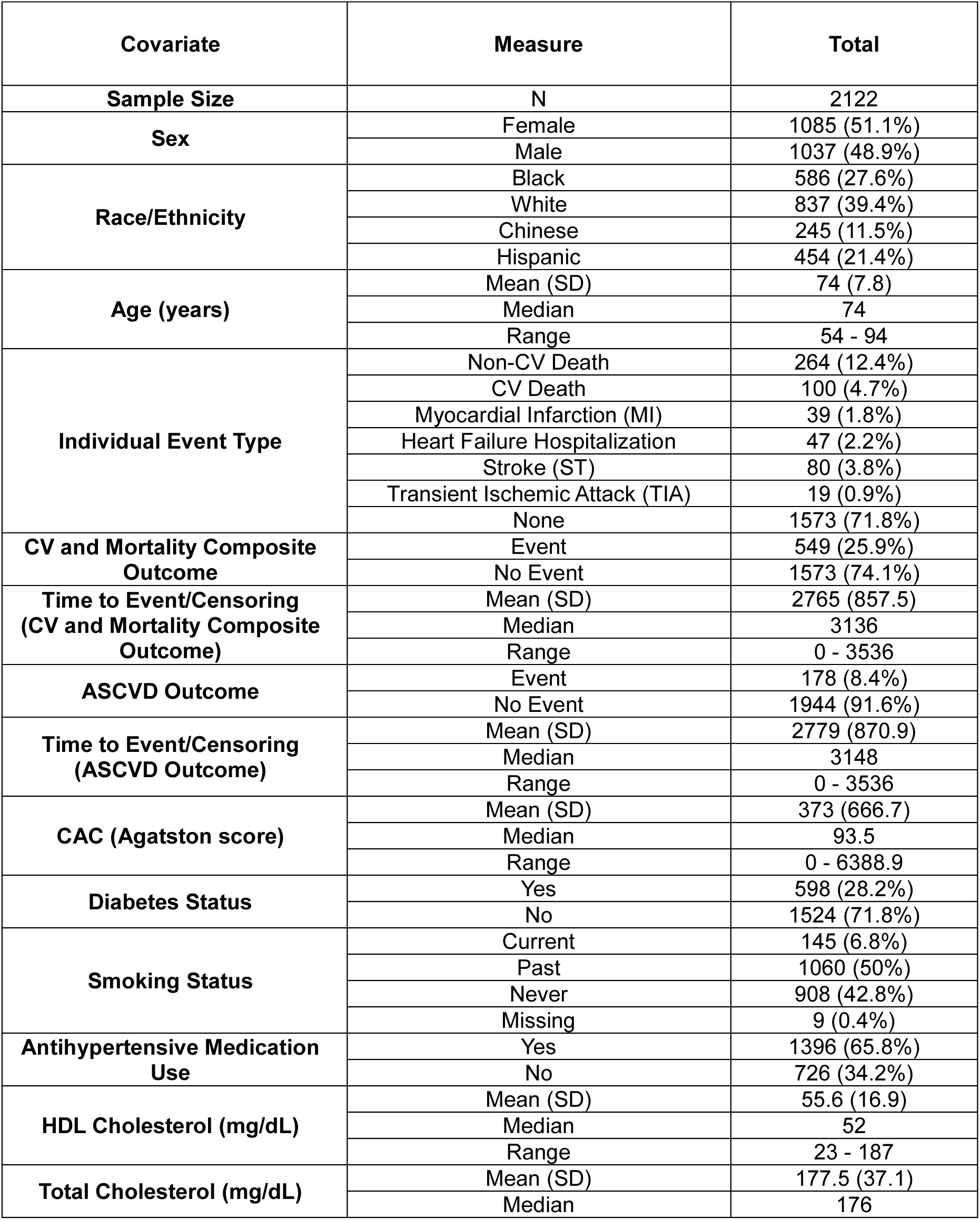
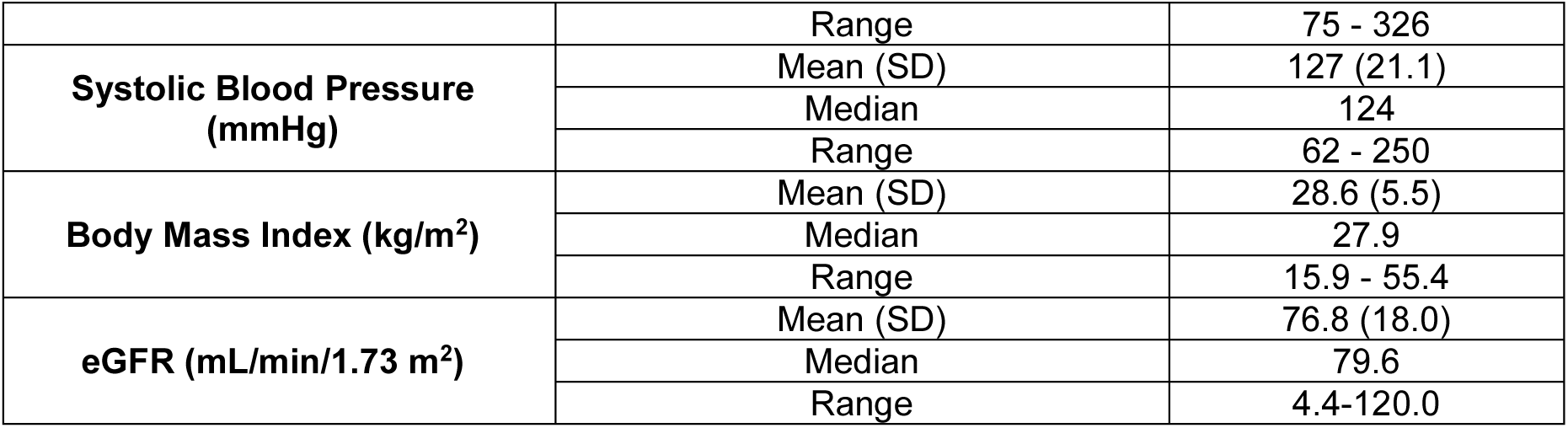
MESA cohort description. Demographic and endpoint table for MESA exam 5 data used for comparative analyses of Prot-CVR and CAC prognostic performance and combined model analyses. Participants included in these analyses were required to have at least one or more known causes of increased CV risk, including individuals with stable CVD, type 2 diabetes, chronic kidney disease, a history of cancer, or > 65 years of age, and available CAC data.

**Table 2:** Measures of test discrimination for Prot-CVR and CAC. Performance metrics: 4-year AUC and C-Index for Prot-CVR and CAC in all participants at exam 5 that are part of the Prot-CVR intended use population (individuals with stable CVD, type 2 diabetes, chronic kidney disease, a history of cancer, or > 65 years of age) and the subset of those participants with non-zero CAC scores. Bold font indicates significant results that have a p-value ≤0.05.

| Group | n | Prot-CVR 4-Year AUC (95% CI) | CAC 4-Year AUC (95% CI) | DeLong's p-value | Prot-CVR C-Index (95% CI) | CAC C-Index (95% CI) | Z-Test p-value |
| --- | --- | --- | --- | --- | --- | --- | --- |
| **CV and Mortality Composite Outcome** |  |  |  |  |  |  |  |
| **All** | 2075 | 0.67<br>(0.63-0.72) | 0.66<br>(0.63-0.7) | 0.649 | **0.68**<br>**(0.65-0.7)** | **0.63**<br>**(0.6-0.65)** | **0.001** |
| **Non-zero CAC** | 1626 | 0.68<br>(0.64-0.72) | 0.64<br>(0.6-0.69) | 0.155 | **0.68**<br>**(0.65-0.7)** | **0.6**<br>**(0.57-0.63)** | **<0.001** |
| **ASCVD Outcome** |  |  |  |  |  |  |  |
| **All** | 1965 | 0.63<br>(0.57-0.69) | 0.67<br>(0.61-0.72) | 0.306 | 0.64<br>(0.60-0.68) | 0.64<br>(0.60-0.68) | 0.721 |
| **Non-zero CAC** | 1531 | 0.64<br>(0.57-0.70) | 0.65<br>(0.58-0.71) | 0.797 | 0.64<br>(0.60-0.67) | 0.61<br>(0.57-0.66) | 0.631 |

Alternatively, assessing predictive performance using the narrower ASCVD outcome, CAC and Prot-CVR performed comparably at both 4 years and over the full 10-year follow-up period. Prot-CVR and CAC both had a C-index of 0.64 (p = 0.721), and a 4-year AUC of 0.63 and 0.67 (p = 0.306), respectively (Table 2). Hazard ratios for the CV and mortality composite and ASCVD outcomes, as well as the individual event types included within the two outcomes, were calculated for Prot-CVR and CAC (Figure 1). Hazard ratios were not significantly different for Prot-CVR and CAC with respect to the ASCVD outcome and the individual event types included within it. The Prot-CVR test had a significantly (p <0.05) larger hazard ratio for the CV and mortality composite outcome and the two individual event types specific to that composite outcome, heart failure hospitalization and all-cause death. Although the primary objective of these analyses was to compare Prot-CVR and CAC risk scores in a heightened-risk population, a small subset of individuals (7.3%) included in the analyses did have a prior cardiovascular event, which falls outside the recommended guidelines for CAC testing. Excluding these individuals resulted in AUC and C-index values that were either unchanged or nominally lower across all evaluated outcomes. These small differences did not change the significance of the comparisons between Prot-CVR or CAC for predicting the composite CV and mortality or ASCVD outcomes (Supplemental Table S3).

**Figure 1:**
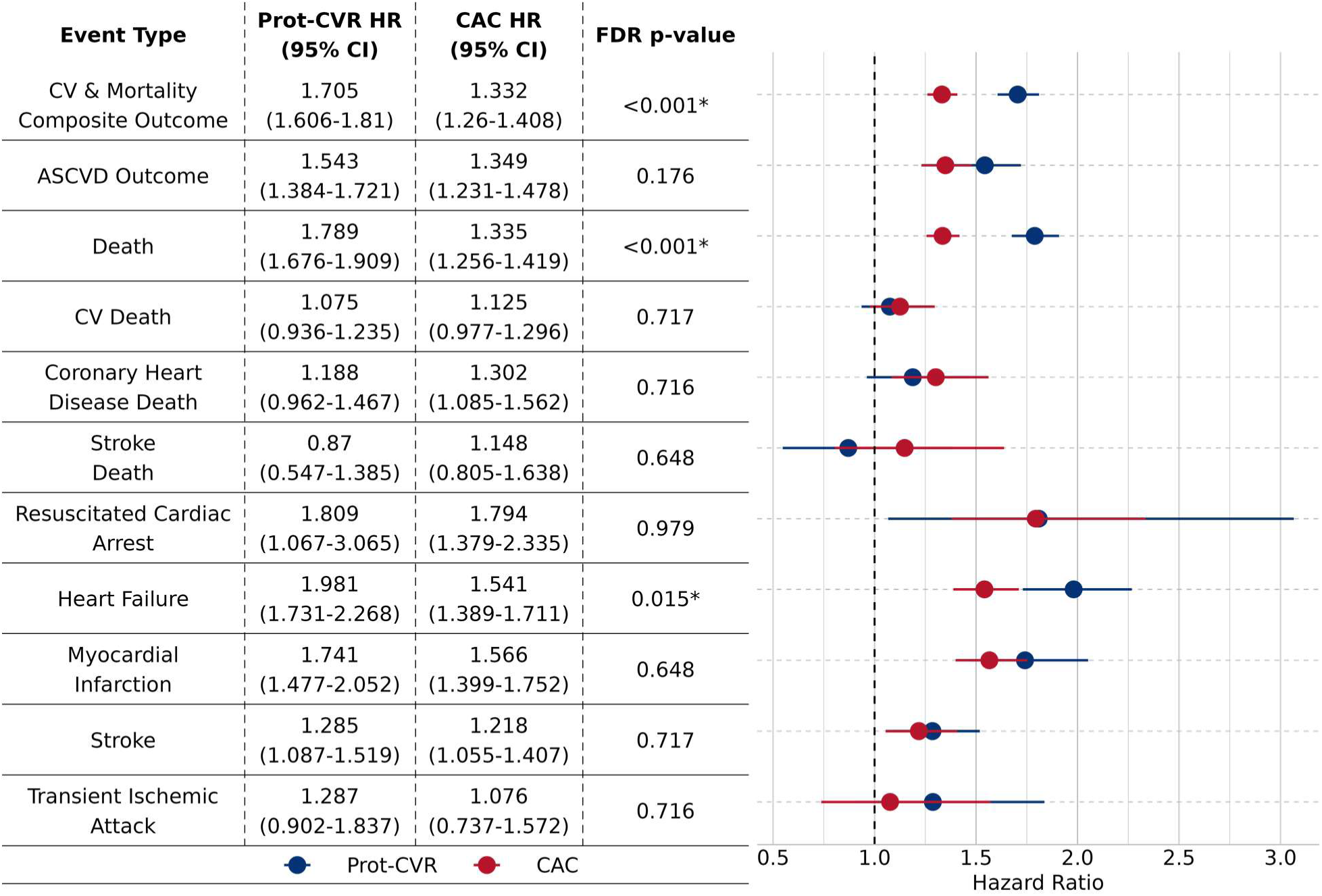
Cox proportional hazard (CoxPH) ratios (HR) by qualifying event type. The CoxPH survival was modeled as event status (“event” or “no event”) to follow-up days (event follow-up days or censoring if no event) distributed by a center scaled metric, Prot-CVR or CAC score. HRs are reported as the exponentiated model coefficients [exp(coeff)], with 95% confidence intervals. HR = 1 indicates no predicted change in risk for the “event” group relative to the “no event” group over the follow-up period, HR >1 indicates increased predicted risk, and HR <1 indicates decreased predicted risk. HRs for Prot-CVR and CAC were compared for each event type using a Z-statistic calculated from the difference in log(HR) divided by the standard error of the difference. Reported p-values were corrected for multiple testing using False Discovery Rate (FDR) adjustment.

In addition to predictive prognostic performance of Prot-CVR and CAC using continuous absolute risk scores, predefined relative risk bins and quartiles were assessed for their ability to accurately stratify participants using the CV and mortality composite outcome. Survival curves stratified by relative risk bins, and quartiles for baseline Prot-CVR and CAC scores are depicted in Figure 2 and Supplemental Figure S2, respectively. Observed 4-year event rates across the 4 risk bins (low, medium-low, medium-high and high for Prot-CVR and no detectable calcium, mild calcification, moderate calcification and extensive calcification) were 3.6%, 7.4%, 13.0%, and 29.1% in Prot-CVR and 4.5%, 6.5%, 8.6%, and 17.1% in CAC, respectively. Relative to the background 4-year event rate of 9.73%, the relative CV risk in these categories was 0.37, 0.76, 1.34, and 2.99 in Prot-CVR, and 0.46, 0.67, 0.90, and 1.76 in CAC, respectively (Supplemental Table S4).

**Figure 2:**
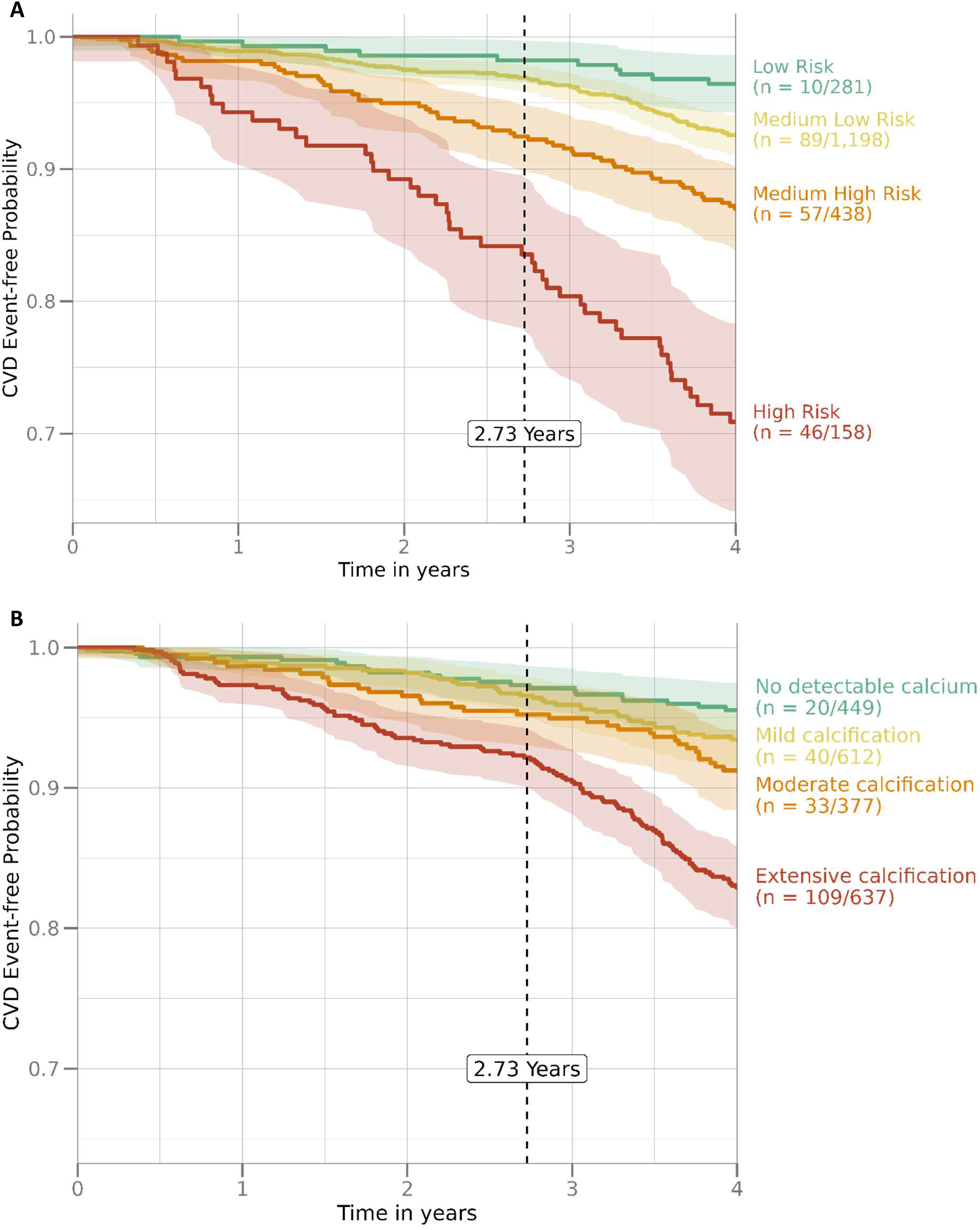
Kaplan-Meier 4-year event-free probability (with 95% CI bands) of all participants stratified by (A) Prot-CVR risk bins, and (B) CAC risk bins. The number of participants diagnosed with a CV or mortality event/total number of participants by risk bin is included in the in-line legend. The median time (years) to CV event across all groups is indicated with a vertical dashed line (2.73 years).

### Prognostic performance of Prot-CVR and CAC combined

Combining Prot-CVR and CAC in a CoxPH model led to improved discrimination in predicting the CV and mortality composite outcome compared to either risk score alone, with a 4-year AUC and C-Index of 0.69 (Figure 3B). When the Prot-CVR risk score was adjusted for traditional risk factors [Diabetes status, sex, high-density lipoprotein cholesterol (mg/dL), systolic blood pressure (mmHg), smoking status, total cholesterol (mg/dL), ethnicity, antihypertensive medication use, and body mass index (BMI)], and for center scaled CAC, prediction was attenuated (HR decreased from 1.71 to 1.48), but Prot-CVR was still significantly associated with event outcomes (Figure 3A). When clinically relevant variables were evaluated alongside the Prot-CVR and CAC metrics as independent predictors of event status in the Cox PH models, age [HR: 1.05 (1.03-1.06); p <0.001], antihypertensive medication use [HR: 1.31 (1.07-1.61); p = 0.0091], sex [female HR: 0.80 (0.67-0.95); p = 0.0117], BMI [HR: 0.98 (0.96-1.00), p = 0.0276], and race/ethnicity [Chinese HR: 0.69 (0.50-0.95); p = 0.0232] were found to be significantly associated with event outcomes (Supplementary Table S5). As Prot-CVR and CAC were center scaled in the Cox modeling, the hazard ratio represents the relative change in hazard for a 1 standard deviation (SD) increase in the scoring metric. A 1 SD increase in the Prot-CVR score is associated with a 48.7% increase in hazard (HR = 1.487), while a 1 SD increase in CAC is associated with a 13.5% increase in hazard (HR = 1.135). Inclusion of age to the combined Prot-CVR + CAC model led to further improvements in predictive performance with a 4-year AUC of 0.72 (versus 0.69 for Prot-CVR+CAC) and C-Index of 0.71 (versus 0.69 for Prot-CVR+CAC).

**Figure 3:**
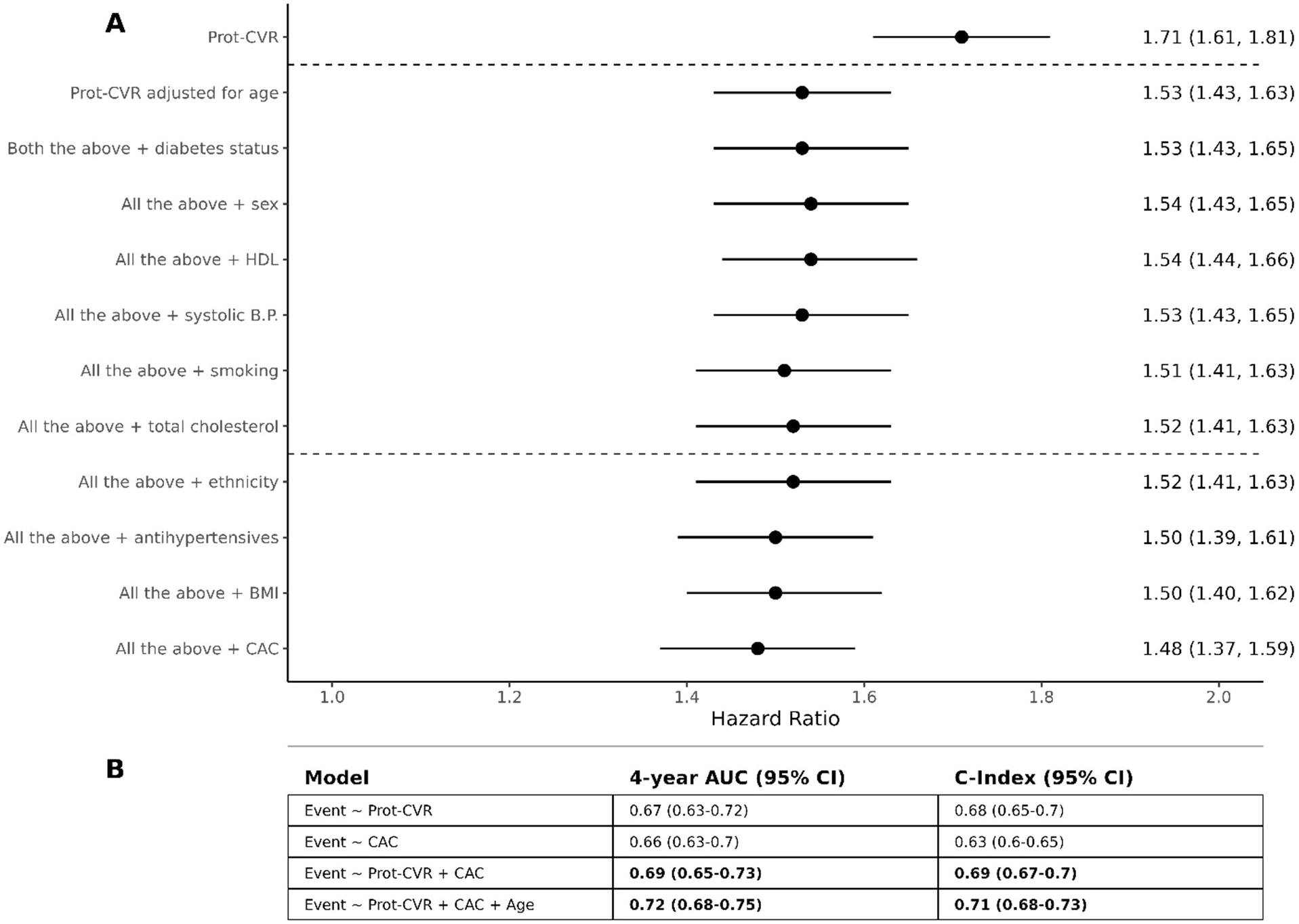
CoxPH models evaluate the performance of one or more scoring metric (Prot-CVR, CAC) with respect to predicting combined cardiovascular events. Models include all participants at exam 5 that are part of the Prot-CVR intended use population (individuals with stable CVD, type 2 diabetes, chronic kidney disease, a history of cancer, or > 65 years of age). **A:** A CoxPH survival model for the composite CV and mortality outcome distributed by center scaled Prot-CVR and adjusted in an add-one manner for clinical covariates (between horizontal dashed lines) known to impact CV risk [diabetes status, sex, high-density lipoprotein cholesterol (mg/dL), systolic blood pressure (mmHg), smoking status, total cholesterol (mg/dL)], for additional clinical covariates significant in a full combined model [ethnicity, antihypertensive medication use, and body mass index (BMI)], and finally for center scaled CAC. **B:** 4-year AUC and C-Index metrics for simplified models. 95% confidence intervals reflect 2-sided confidence interval around derived 4-year AUC and C-Index metrics. Bold font indicates a significant (p ≤0.05) increase in performance of the simplified combined models (Prot-CVR + CAC and Prot-CVR + CAC + Age) relative to Prot-CVR and/or CAC only models, based on a two-sided Delong’s test (AUC) and C-Index Z-score.

To further assess the complementarity of Prot-CVR and CAC, a sub cohort of individuals with CAC = 0 at exam 1 were evaluated to determine if Prot-CVR scores could provide additional information for short- and long-term event risk, using the CV and mortality composite outcome. Mean Prot-CVR scores in the 4-year “event” group were significantly higher (mean Prot-CVR: 0.232; p <0.01) than in the “no event” group (mean Prot-CVR: 0.148), as shown in Figure 4. Similarly, when looking at the full follow-up period, mean Prot-CVR scores were significantly higher in the “event” group (mean Prot-CVR: 0.180 versus 0.130, in the event and “no event” group, respectively). Using the high-risk bin threshold (a Prot-CVR score over 0.5) we were able to identify a subset of individuals with CAC = 0, and a 5.21-fold relative increased risk of having a CV event within 4 years (Supplementary Table S6).

**Figure 4:**
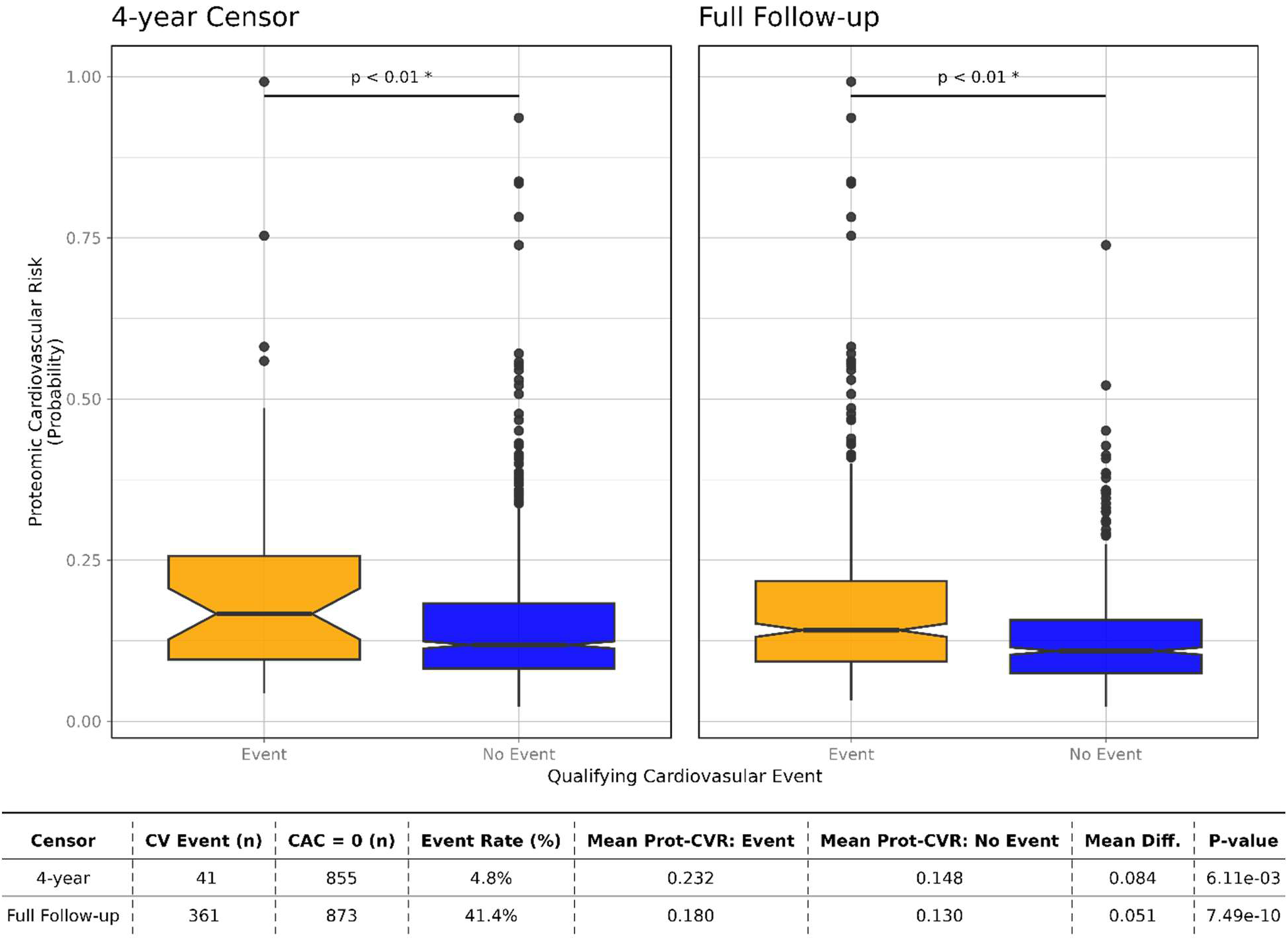
Prot-CVR scores for participants with a CAC score of 0 at Exam 1 for CV and mortality composite outcome events at 4 years and over the full follow-up period, shown as a notched box plot with outliers. Differences between mean Prot-CVR scores in the “event” versus “no event” group were assessed using a one-sided t-test with an alternative hypothesis that the mean “event” score would greater than the “no event” score. Group means between “event” and “no event” groups that are significantly different from each other (p-value <0.01) are indicated with “*”. Non-CV mortality rate was 2.2% over 4 years and 23.4% over the full follow-up period.

### Sensitivity to detect an increase in predicted risk in advance of an upcoming CV outcome event

Prot-CVR and CAC scores were significantly higher (Prot-CVR: 0.15 versus 0.11, and CAC: 197.13 versus 97.03; p <0.001) at exam 1 (baseline) in individuals who had events (using the CV and mortality composite outcome) after exam 5 compared to those that did not have an event during the full 10-year follow-up period (Supplemental Table S7). Both Prot-CVR and CAC scores also increased significantly more from exam 1 to exam 5 in those who had an event after exam 5 compared to the “no event” group (Figure 5, Table 3). Prot-CVR scores increased 15.3% from exam 1 to exam 5 in the “event” group compared to only 7.0% in the “no event” group (p <0.001), and CAC scores increased 325.9 Agatston units in the “event” group compared to 180.0 Agatston units in the “no event” group (p <0.001).

**Figure 5:**
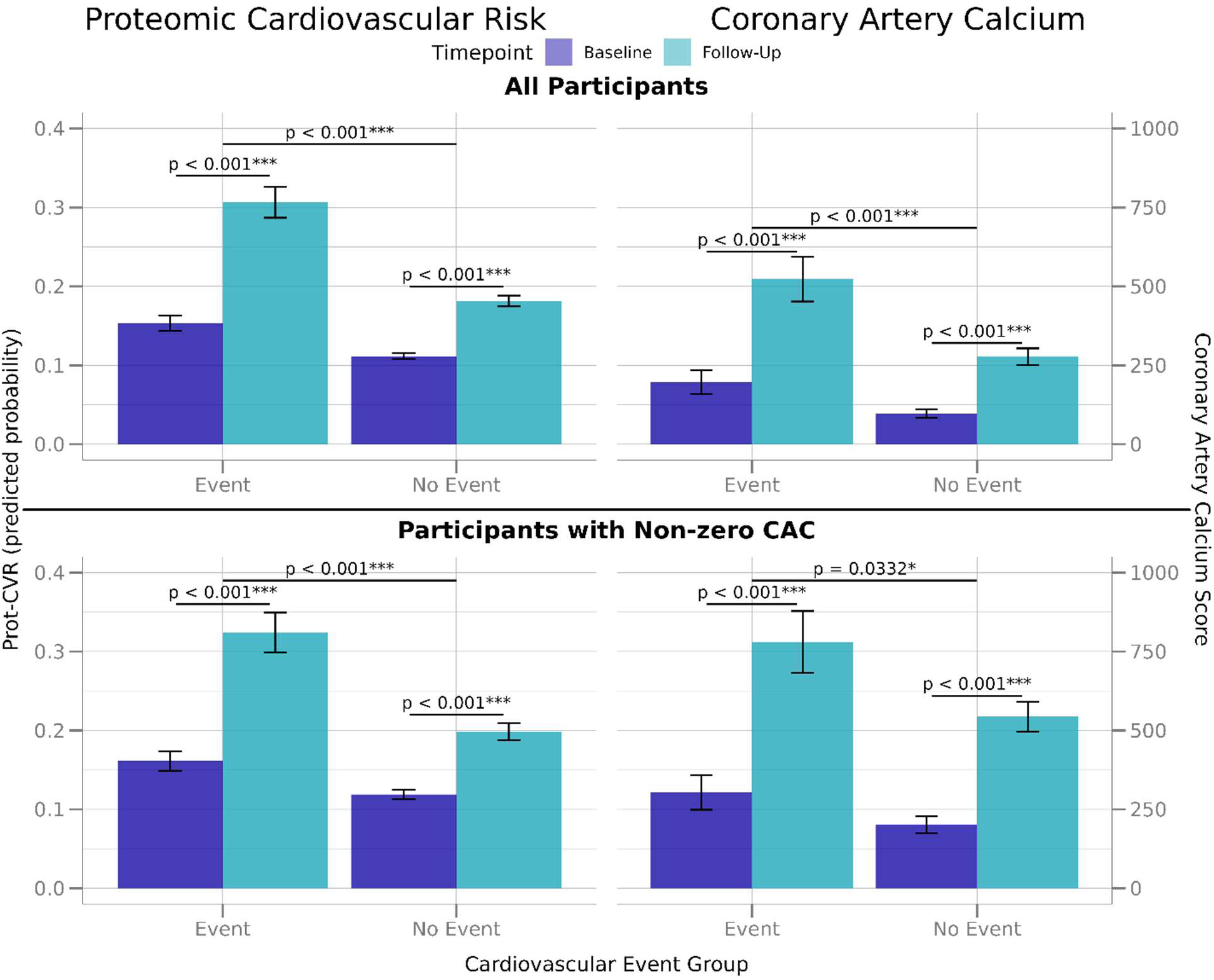
Sensitivity to change in risk as measured by Prot-CVR and CAC from baseline to follow-up (Exam 5) in advance of an upcoming CV event. Change in risk from baseline to exam 5 in participants with paired samples that are part of the Prot-CVR intended use population (individuals with stable CVD, type 2 diabetes, chronic kidney disease, a history of cancer, or over 65 years old) that went on to have an event (CV and mortality composite outcome: MI, Stroke/TIA, HF hospitalization or all-cause death). Error bars represent 95% confidence intervals around the mean score. The left panels represent Prot-CVR predictions, and the right panels are CAC scores. The top panels are for all participants, while the bottom panels represent participants with non-zero CAC scores at exam 1 and exam 5. Significance testing for within-group change from baseline to follow-up and between-group difference was calculated using two-sided, two-sample t-tests (alpha = 0.05), *** <0.001, ** <0.01, * <0.05.

**Table 3:**
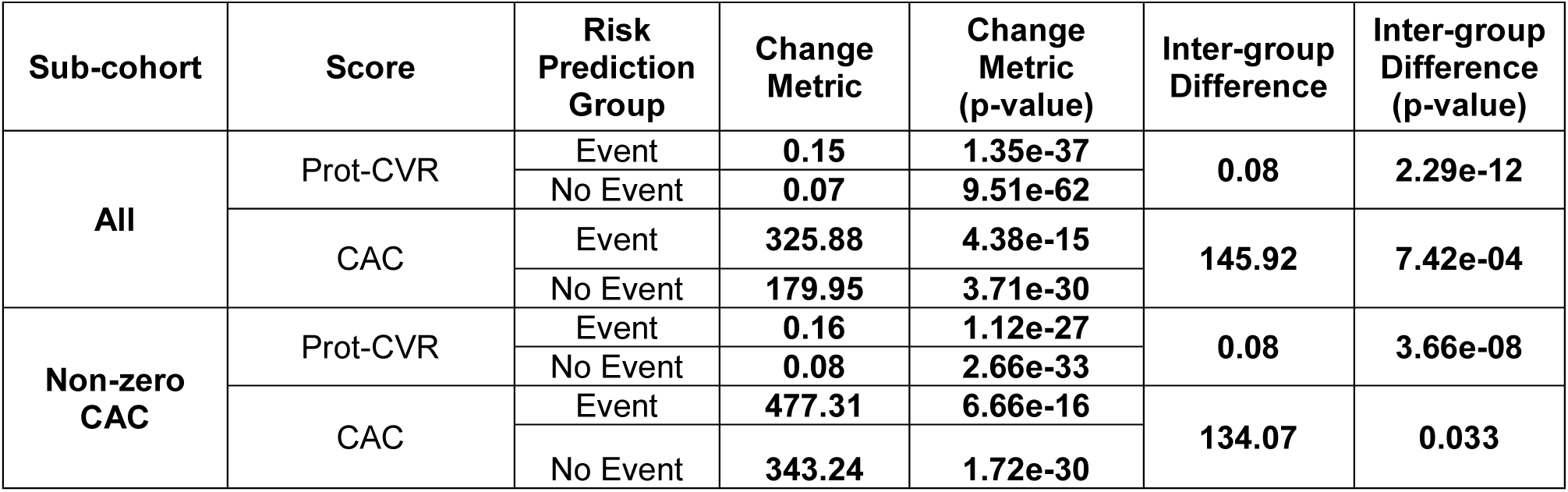
Prot-CVR and CAC changes in risk in advance of an upcoming CV event. Change in risk from baseline to exam 5 in participants with paired samples that are part of the Prot-CVR intended use population (individuals with stable CVD, type 2 diabetes, chronic kidney disease, a history of cancer, or over 65 years old) that went on to have an event (CV and mortality composite outcome: MI, Stroke/TIA, HF hospitalization or all-cause death) after exam 5 versus participants that remained event free during follow-up. Non-zero CAC sub-cohort is comprised of participants with CAC >0 at both exam 1 and exam 5. The change metric was calculated as the mean follow-up value minus the mean baseline value for each risk prediction group. The inter-group difference is the difference in the change metric between the risk prediction groups (“Event” and “No Event”) for each metric. T-tests are 2-tailed. Bold font indicates a significant result (p ≤0.05).

## Discussion

In this study of MESA participants with risk factors indicative of increased cardiovascular event risk, we found that CAC was comparable to the Prot-CVR test in informing the risk of a broad range of events, including coronary events, strokes/TIAs, heart failure and all-cause deaths within a 4-year time horizon. However, when assessing prognostic performance over a full 10-year follow-up period (C-index), the Prot-CVR test performance improved, while CAC performance diminished. These results suggest that Prot-CVR can better risk stratify long-term outcomes, however, it is possible or even likely that the improved performance is a result of the higher event rate (25.9% 10-year event rate versus 9.73% 4-year event rate) and Prot-CVR may be better at up-classifying risk. These results are consistent with previous findings^14^ and further supported by our subset analyses showing Prot-CVR’s risk assessment performance improves in the subset of individuals with non-zero CAC (which had a higher overall event rate of 29%) and by the stratification of individuals using Prot-CVR relative risk bins (which showed a higher event rate in the high-risk group compared to CAC).

When assessing hazard ratios for composite and individual event types, the Prot-CVR test had a significantly larger hazard ratio for the CV and mortality composite outcome indicative of its broader dynamic range and for the two individual event types specific to that composite outcome, heart failure hospitalization and all-cause death. These results were as expected since Prot-CVR was developed to predict these additional outcomes.

Next, we assessed whether the Prot-CVR test, in combination with CAC and other traditional clinical risk factors, could enhance prognostic performance beyond what is achievable through these measures individually. Combining Prot-CVR and CAC improved discrimination compared to either risk score alone in predicting the CV and mortality composite outcome. Additionally, even after adjusting for other clinical risk factors that were found to be significantly associated with event outcomes (age, antihypertensive medication use, gender, BMI, and ethnicity) both Prot-CVR and CAC scores were significantly associated with outcomes. Together, these results suggest that Prot-CVR and CAC provide novel information about CV risk and may be complementary tools to improve and refine risk assessments.

Although a CAC score of 0 is associated with a relatively low short-term risk of cardiovascular events, it does not eliminate risk. In our cohort of individuals with a CAC score of 0, CV or mortality events occurred in 4.8% (2.6% were ASCVD events) of individuals over 4 years and increased to 41.4% (18% were ASCVD events) over the full follow-up period. Therefore, we evaluated Prot-CVR in a sub cohort of individuals with CAC = 0 at exam 1 to determine if Prot-CVR scores could help identify participants at risk of an event. Prot-CVR score mean values in the “event” group were higher (p <0.001) than in the “no event” group using 4-year censoring and the full follow-up period. Additionally, utilizing the high-risk bin threshold, we were able to identify a subset of individuals with a 5.21-fold elevated relative risk of a CV event or mortality within the next 4 years. These results suggest that using Prot-CVR in combination with CAC scores may provide better risk stratification and help identify participants with zero calcium who are at risk for a CV or mortality event.

Lastly, we assessed longitudinal performance of Prot-CVR and CAC scores for their sensitivity to detect a change in risk in advance of an upcoming CV or mortality event. Absolute risk scores significantly increased in both Prot-CVR and CAC in participants approaching an event (after exam 5) versus those that remained event free during the full 10-year follow-up period. Subset analyses in participants with non-zero CAC revealed similar results, although the p-value for change in CAC scores between the “event” and “no event” group declined (p-value <0.001 versus 0.0332). This finding may reflect a known limitation of CAC serial testing in individuals taking plaque-stabilizing drugs such as statins, where CAC score often increases with treatment^13^, and statin use was higher in the subset of individuals with non-zero CAC. Overall, both CAC and Prot-CVR scores increased more in participants who went on to have a CV event, compared to those who remained event free, however, Prot-CVR may be better suited for longitudinal assessment in individuals with non-zero CAC since CAC predictive performance declined over time in this subset. Additionally, these individuals are also more likely to be on cardioprotective drugs, and Prot-CVR has previously been shown to be sensitive to changes in CV risk in response to pharmacological interventions, including novel cardioprotective drugs^24, 25^.

This study has many strengths and some potential limitations. While MESA is a well-characterized, contemporary, racially diverse cohort, the populations used for our analyses differ from previous studies. Notably, these analyses were run in participants that met the intended use population for the Prot-CVR test those having ≥1 high CV risk factors. We did not evaluate whether Prot-CVR would have an identical role as CAC in borderline or intermediate risk patients. However, a subset analysis excluding participants with prior cardiovascular events at exam 5, cases considered in-scope for the intended use population of the Prot-CVR test, but out of the recommended guidelines for CAC did not change the significance of the comparisons between Prot-CVR and CAC performance metrics. Similarly, although analyses were run in participants that met intended use criteria for Prot-CVR, the event rates in this MESA dataset were significantly lower than the event rates in the 22,849 participants used to train and validate the Prot-CVR. Another limitation is that we evaluated CAC for prediction of a broad composite endpoint rather than coronary events exclusively. The rationale for using the broad composite endpoint was based on the potential of these tools to allocate pharmacotherapies, as the adverse outcomes that would be modulated by these prognostic tests (e.g., GLP-1 receptor agonists, SGLT2 inhibitors, anti-hypertensives) are not limited to the reduction of myocardial events. This concern was addressed by the same assessment of prognostic performance in ASCVD outcomes alone (MI, resuscitated cardiac arrest, stroke, CHD death, or stroke death) and in individual event types.

In summary, these findings suggest that prognostic protein scoring, such as Prot-CVR, can provide a complementary tool for CV risk assessment and that Prot-CVR and CAC together may provide a more comprehensive assessment of patient risk and aid in medical management and monitoring. However, the improvement in the C-index with addition of Prot-CVR to CAC is modest. Further studies will have to establish whether it is clinically meaningful and cost-effective. Currently, an attractive application of Prot-CVR may be to resolve which patients with CAC = 0 are at heightened risk of cardiovascular events.

## Data Availability

MESA (Multi-Ethnic Study of Atherosclerosis) data are accessible through the NHLBI Biologic Specimen and Data Repository Information Coordinating Center (BioLINCC). Researchers must submit a proposal for data use, ensuring compliance with HIPAA regulations and participant consent, with data available in both public and restricted formats via dbGaP.

## Acknowledgments

The authors are grateful to the MESA study participants, study teams, physicians, and medical staff for their valuable contributions. A full list of participating MESA investigators and institutions can be found at http://www.mesa-nhlbi.org. The authors also thank the SomaScan Assay team, platform bioinformatics team for quality control, data management team for clinical data organization, and Jessica Williams leading the execution of the collaboration agreement with the study institution.

## Sources of Funding

This study was in part supported by a NIH grant award 1R01HL159081 to P. Ganz. This research was also supported by contracts 75N92025D00022, 75N92020D00001, HHSN268201500003I, N01-HC-95159, 75N92025D00026, 75N92020D00005, N01-HC-95160, 75N92020D00002, N01-HC-95161, 75N92025D00024, 75N92020D00003, N01-HC-95162, 75N92025D00027, 75N92020D00006, N01-HC-95163, 75N92025D00025, 75N92020D00004, N01-HC-95164, 75N92025D00028, 75N92020D00007, N01-HC-95165, N01-HC-95166, N01-HC-95167, N01-HC-95168 and N01-HC-95169 from the National Heart, Lung, and Blood Institute, and by grants UL1-TR-000040, UL1-TR-001079, UL1-TR-001420, UL1TR001881, and R01HL105756 from the National Center for Advancing Translational Sciences (NCATS). The authors thank the other investigators, the staff, and the participants of the MESA study for their valuable contributions. A full list of participating MESA investigators and institutions can be found at http://www.mesa-nhlbi.org.

This publication was also developed under the Science to Achieve Results (STAR) research assistance agreements, No. RD831697 (MESA Air) and RD-83830001 (MESA Air Next Stage), awarded by the U.S Environmental Protection Agency (EPA). It has not been formally reviewed by the EPA. The views expressed in this document are solely those of the authors, and the EPA does not endorse any products or commercial services mentioned in this publication.

This paper has been reviewed and approved by the MESA Publications and Presentations Committee.

## Abbreviations

CAC: coronary artery calcium
ASCVD: atherosclerotic cardiovascular disease
CV: cardiovascular
CoxPH: Cox Proportional Hazard
Prot-CVR: proteomic cardiovascular risk
MESA: Multiethnic Study of Atherosclerosis
MI: myocardial infarction
HF: heart failure
CVD: cardiovascular disease
TIA: transient ischemic attack
CHD: coronary heart disease
ROC: Receiver Operating Characteristic
AUC: Area Under the Curve
C-Index: Concordance-Index
KM: Kaplan-Meier
FDR: False Discovery Rate
HR: hazard ratio

